# Gut unclassified Ruminococcaceae Reweights Cortical Functional Gradients and Small-World Topology with Links to Mood and Diet

**DOI:** 10.1101/2025.10.31.25338831

**Authors:** Xiaobo Liu, Zhengyuan Huang, Lang Liu, Sanwang Wang, Xiaoqiang Liu

## Abstract

The microbiota-gut-brain axis is a key conduit linking metabolism, mood, and cognition; however, its position within the continuous functional hierarchy of the cortex and the underlying mechanisms are unclear. In this study, a cross-modal brain-gut dataset from 88 healthy male participants was utilized. By integrating functional magnetic resonance imaging (fMRI) gradient analysis, microbiome sequencing, and dietary behavioral information, this study systematically evaluated the relationships between the abundance of *unclassified Ruminococcaceae* (*Ruminococcaceae_unc*), cerebral functional hierarchy, network topology, and emotional symptoms. The results demonstrated that increased abundance of this bacterial group drives a functional shift in the brain from unimodal to transmodal hubs, accompanied by a drift of the small-world network toward randomization. Functional gradient values were significantly negatively correlated with depression and anxiety scores and were tightly coupled with latent components in the dietary behavioral dimension, including education, physical activity, and nutrient intake. Transcriptomic analysis further revealed that the GPCR-Rho/integrin-vesicular trafficking pathway may serve as the key molecular mechanism. In conclusion, this study proposes a multiscale coupling framework encompassing the gut microbiota, functional gradients, and emotional health, thereby providing a theoretical basis for the development of microbiota-targeted intervention strategies for modulating transmodal emotions and cognition.

## 1. Introduction

In recent years, the microbiota-gut-brain axis has been regarded as a multilevel signaling pathway that connects the metabolic, neural, and immune systems. It provides a novel theoretical framework for understanding the regulation of emotion, cognition, and the autonomic nervous system[1,2]. A multimodal cross-sectional study involving healthy adult males revealed that both indices of autonomic nervous system homeostasis and the functional connectivity strength of the central executive network were positively correlated with the relative abundance of certain *unclassified Ruminococcaceae* (*Ruminococcaceae_unc*). Furthermore, a significant interaction effect was observed between autonomic nervous system function and the central executive network, in which dietary fiber intake exerted a partial mediating role[3]. This finding suggests that the coupling between autonomic nervous system balance and the high-level cognitive control network depends on the colonization level of specific gut microbiota. However, most studies of this type still rely on discrete brain network metrics and have not yet addressed key questions, including the precise localization of the associations between microbiota and neural activity within the continuous hierarchy of the cerebral cortex, the directionality of these associations, and the significance of such associations in characterizing overall brain function. Meanwhile, there remains a lack of systematic descriptions of the regulatory effects of daily dietary structure and anxiety-depressive spectrum states on this axial pathway.

From a macroscopic perspective, brain function can be characterized using two complementary frameworks. The first is the high-dimensional topological perspective, which divides the cerebral cortex into several discrete functional networks, such as the default mode, central executive, and salience networks, and emphasizes the graph-theoretic properties of nodes and edges[4]. The second is the low-dimensional continuous representation, namely, the functional gradient. This method maps whole-brain connectivity patterns into smooth coordinate axes via dimensionality reduction algorithms: the first functional gradient accounts for the largest variance along the direction from "sensorimotor regions to transmodal association regions,” while the subsequent second and third gradients supplement dimensions such as the anterior-posterior and temporal-parietal axes[5]. Compared with traditional graph-theoretic methods, functional gradients not only preserve information within and between networks but also enable the quantification of the relative positions of different cortical regions in the hierarchical spectrum. Consequently, they provide a more advantageous coordinate system for exploring the connections between local metabolism and whole-brain function[6].

In recent years, integrated studies of neuroimaging and transcriptomics have further revealed that brain functional gradients are highly coupled with the spatial distribution of multiple types of neurotransmitter receptors and their encoding genes across the cerebral cortex[7,8]. Along the first functional gradient, progressing from the unimodal to transmodal regions, the density of multiple receptors coupled to signal transduction, including serotonin, dopamine, glutamate, gamma-aminobutyric acid (GABA), and cholinergic receptors, gradually increases. In contrast, the sensorimotor end is enriched with voltage-gated ion channels and axon guidance molecules[9,10]. These neurotransmitter systems regulate dendritic morphology and synaptic plasticity through small-molecule signaling cascades, and their gene co-expression modules are regarded as transcriptional topographic maps that link microscale molecular processes to macroscale functional structures[11]. Therefore, analyzing the distribution of receptors and genes within the coordinate system of functional gradients not only provides molecular clues for understanding the hierarchical information processing of the cerebral cortex but also lays a foundation for exploring how gut-derived metabolites (such as butyrate and trimethylamine N-oxide) regulate the trans-network dynamic balance through specific receptor clusters, thereby bridging the gap between macroscopic functions and cellular-molecular mechanisms.

Against this backdrop, based on existing data, this study systematically evaluated the following research questions using a functional gradient approach: First, to delineate the coupling relationships between the abundance of *Ruminococcaceae_unc* and functional gradients at both spatial and individual levels; Second, to examine the association structure between core metrics, including transmodal and unimodal gradient values, and depression, anxiety, as well as essential lifestyle and nutritional factors; Third, to establish a joint model of gradient metrics and small-world topological parameters, thereby dissecting the remodeling mechanism underlying the transition of brain networks from randomization to high efficiency driven by the abundance of specific gut microbiota;Fourth, to identify the molecular mechanisms of gradient remodeling through enrichment analysis of receptors and synaptic molecular pathways.

This integrated framework provides a novel theoretical basis for understanding functional gradient plasticity and developing transmodal interventions targeting specific gut microbiota. Elucidating these questions will advance our understanding of the hierarchical neurobiological mechanisms underlying the microbiota–gut–brain axis and provide a theoretical framework for developing intervention strategies based on functional gradient plasticity and microbiota modulation, such as biofeedback, targeted prebiotics, and metabolic inhibitors.

## 2. Results

### 2.1 Primary Functional Gradient

This study utilized the cross-modal brain-gut dataset publicly available from Miranda-Angulo et al. (2024) on OpenNeuro (accession number: ds004648) and SRA (accession number: PRJNA1000574), which included 88 healthy Colombian males aged 21–40 years. For all participants, simultaneous data collection was performed, including 10-minute 3T resting-state functional magnetic resonance imaging (fMRI) (analyzed using the Yeo-17 network atlas and Schaefer-400 parcellation), fecal 16S rRNA (V3–V4 region) microbiome sequencing, and a food frequency questionnaire. This dataset provides a unified and high-quality foundation for exploring the interactions between functional connectivity, gut microbiota, and diet.

The first primary functional gradient (FG-1), derived by applying diffusion mapping to the group-averaged resting-state functional connectivity matrix, explained 25.2% of the variance in the functional connectivity. Consistent with previous literature, the gradient direction was defined such that "negative values correspond to unimodal cortices and positive values correspond to transmodal cortices." FG-1 exhibited a canonical "sensory-transmodal" continuum: the primary visual cortex, precentral and postcentral gyri (somatomotor regions), and superior temporal gyrus (auditory region) were localized at the negative end, while transmodal hubs, including the medial prefrontal cortex, angular gyrus, posterior cingulate cortex-precuneus, ventrolateral-ventromedial prefrontal cortex, and parahippocampal gyrus, were localized at the positive end (Fig. 1A).

**Figure 1.**
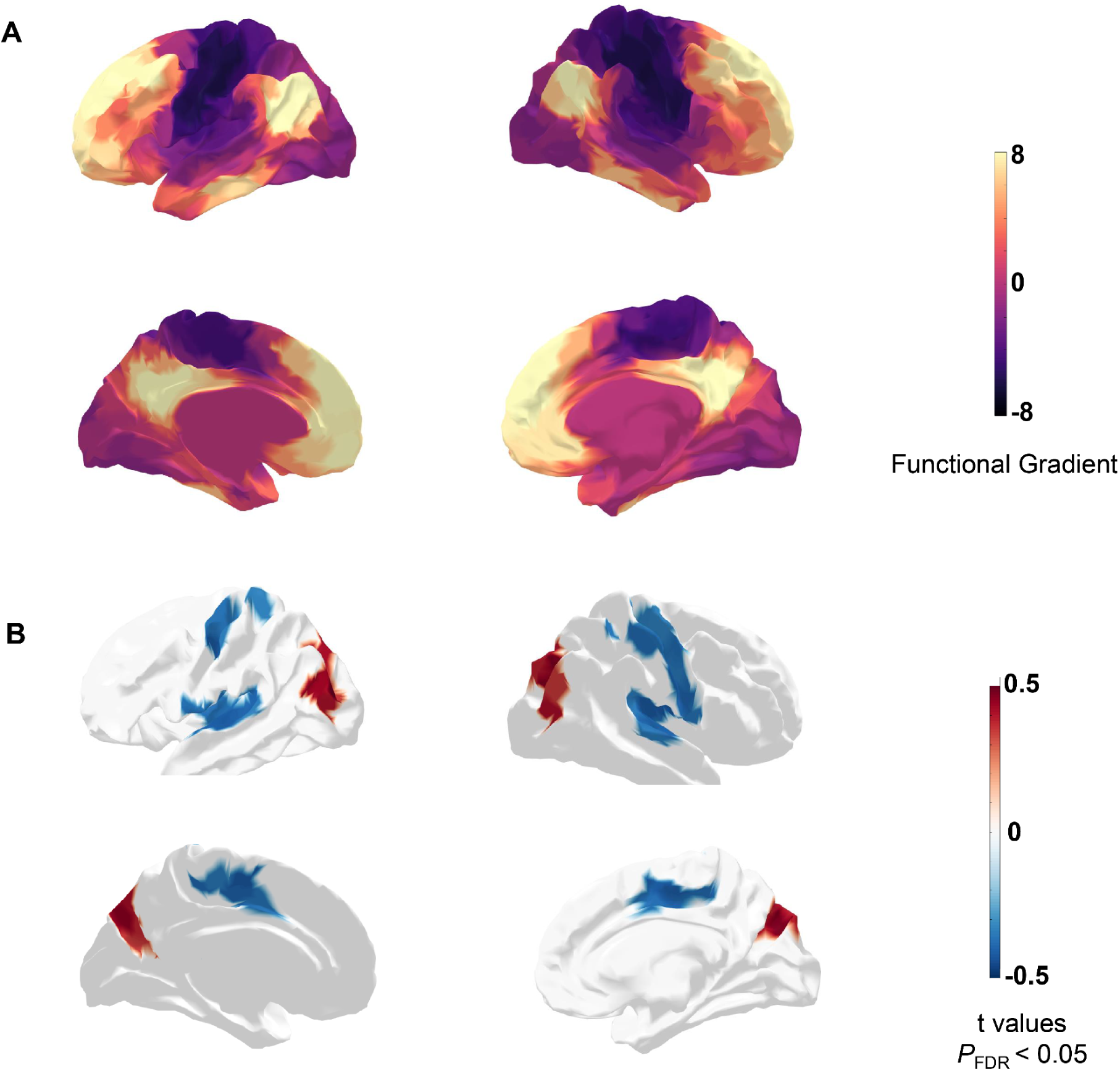
Functional gradient and its correlation with *Ruminococcaceae_unc*. (A) Cortical functional gradient maps of the left and right hemispheres in the lateral and medial views. The color scale reflects the distribution of functional gradients from sensory/motor regions (purple) to higher-order transmodal regions (yellow).(B) Brain regions showing significant correlations between functional gradient values and relative abundance of *Ruminococcaceae_unc*. Red and blue denote positive and negative correlations, respectively (two-tailed test, FDR-corrected, *p* < 0.05).

After controlling for age, sex, and head motion, a partial correlation analysis was conducted between vertex-wise FG-1 values and the relative abundance of *Ruminococcaceae_unc* (FR_*Rumi_unc*). Vertex-level thresholding with pFDR < 0.001 and cluster-level FDR correction (pFDR < 0.05) were applied. The results showed that Higher FR_*Rumi_unc* abundance was significantly associated with increased FG-1 weights in the bilateral angular gyrus (Brodmann Area 39; peak r = 0.34, pFDR = 0.003) and lateral associative visual cortex (Brodmann Area 19; r = 0.31, pFDR = 0.006). In contrast, decreased FG-1 weights were observed in the primary motor cortex (Brodmann Area 4), somatosensory association cortex (Brodmann Area 5), supramarginal gyrus/inferior parietal lobule (Brodmann Area 40), gustatory cortex (Brodmann Area 43), and anterior cingulate cortex (Brodmann Area 24) (-0.33 ≤ r ≤ -0.26, all pFDR < 0.015; Fig. 1B).

Overall, the abundance of FR_*Rumi_unc* was closely associated with the reweighting of the macroscopic functional hierarchy, suggesting that the gut microbiota may shape the hierarchical shift of the brain from unimodal to transmodal cortices.

### 2.2 Small-World Properties of Networks

After confirming that FG-1 was reweighted by microbial abundance, we examined whether this hierarchical rearrangement was accompanied by changes in the topology of the whole-brain functional network. Based on individual functional connectivity matrices under the primary sparsity threshold, small-world properties were calculated, and a correlation analysis was performed between these properties and the relative abundance of FR_*Rumi_unc* (Fig. 2A).

**Figure 2.**
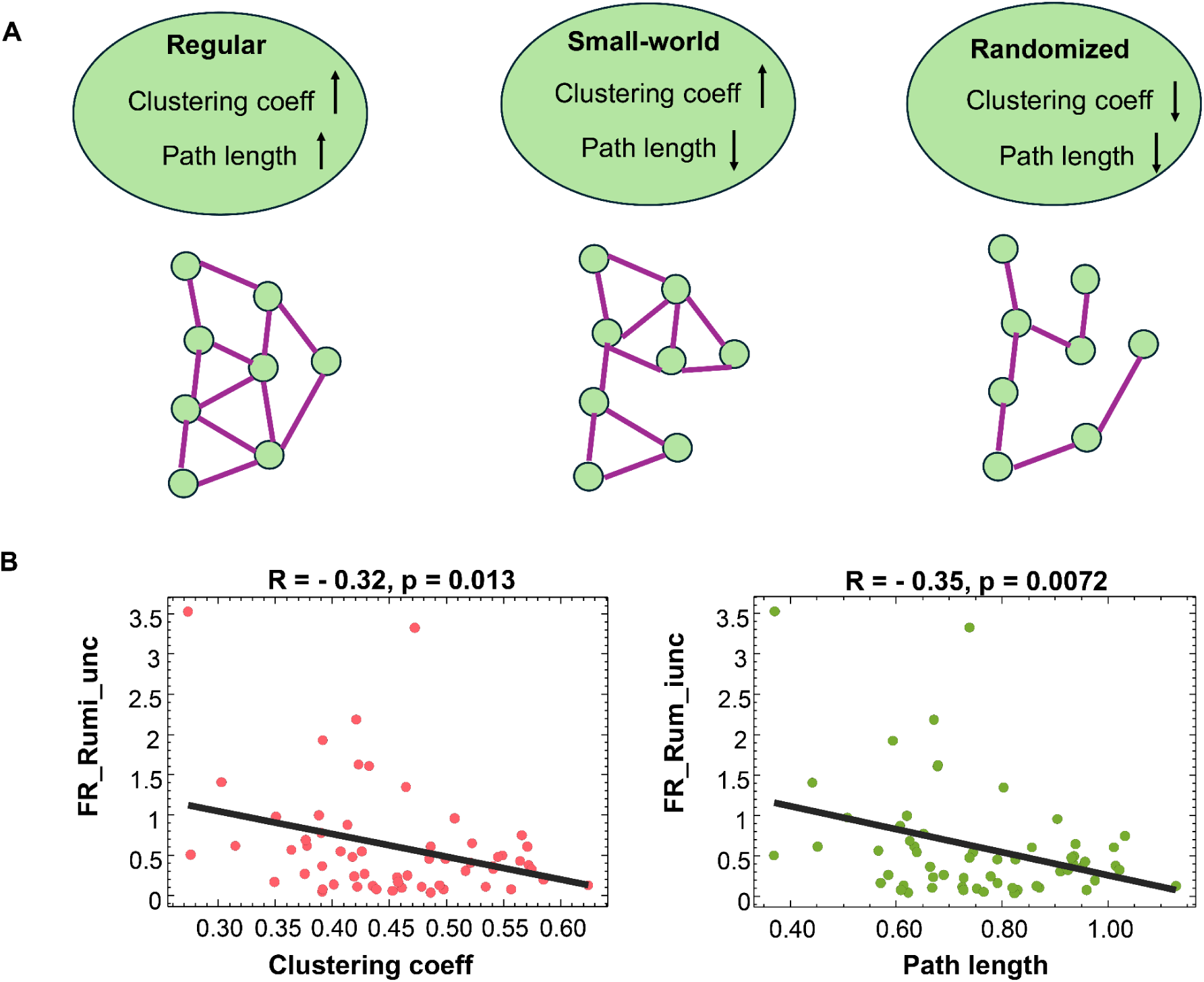
Network topology and its correlation with *Ruminococcaceae_unc*. (A) Schematic illustration of network organizations: regular networks (high clustering coefficient, long path length), small-world networks (high clustering coefficient, short path length), and randomized networks (low clustering coefficient, short path length).(B) Scatter plots showing significant negative correlations between the relative abundance of *Ruminococcaceae_unc* and network topological properties. *Ruminococcaceae_unc* abundance was negatively correlated with the clustering coefficient (left, R = –0.32, *p* = 0.013) and path length (right, R = –0.35, *p* = 0.0072).

The results showed that microbial abundance was significantly negatively correlated with the global clustering coefficient (C) (r = –0.32, p = 0.013; Fig. 2B, left) and also negatively correlated with the characteristic path length (L) (r = –0.35, p = 0.0072; Fig. 2B, right). In other words, as the abundance of FR_*Rumi_unc* increased, the functional network gradually deviated from the canonical small-world structure and drifted toward a "randomized" topology characterized by higher global efficiency and lower local specialization.

This finding indicates that the gut microbiota can not only remodel the hierarchical shift of the brain from unimodal to transmodal cortices but also alter the macroscopic balance of the brain along the integration-segregation dimension.

### 2.3 Correlation Between Functional Gradient Differences and Depression-Anxiety Symptoms

After confirming that network topology drifts with microbial abundance, we investigated its clinical implications. We extracted the FG-1 weight values of brain regions most correlated with microbial abundance and conducted a correlation analysis between these values and depression/anxiety scores, controlling for age, sex, and head motion.

The results showed that functional gradient values were significantly negatively correlated with depression scores (R = –0.27, p = 0.036) and also significantly negatively correlated with anxiety scores (R = –0.31, p = 0.019; Fig. 3A). Further results on cortical distribution indicated that the brain regions with significant correlations were mainly located in transmodal integration hubs (Fig. 3B)

**Figure 3.**
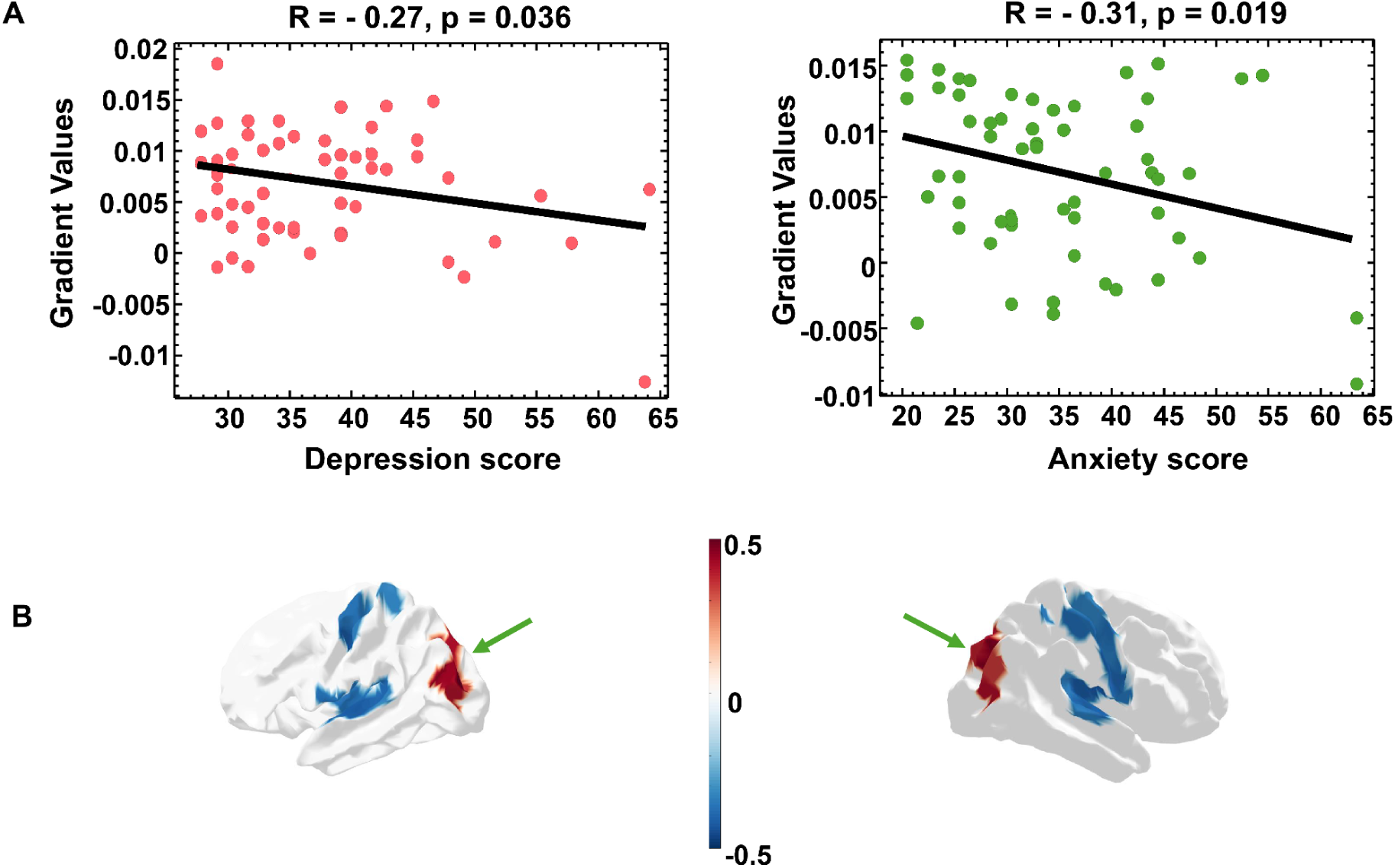
Associations between functional gradient and emotional symptoms. (A) Scatter plots showing negative correlations between functional gradient values and clinical symptom scores. Gradient values were negatively correlated with depression scores (left, R = –0.27, *p* = 0.036) and anxiety scores (right, R = –0.31, *p* = 0.019).(B) Brain surface maps highlighting the cortical regions showing significant associations between gradient values and depression/anxiety symptoms (FDR-corrected, *p* < 0.05). Red and blue indicate positive and negative correlations, respectively. Green arrows indicate significant clusters.

In summary, these results suggest that the more severe the emotional symptoms, the more an individual’s functional gradient tends toward the unimodal sensory-motor end, and the relative contribution of transmodal integration hubs decreases. This finding reinforces the potential link between the "gut microbiota-driven reweighting of functional hierarchy" and emotional vulnerability.

### 2.4 Integrated Analysis of Dietary Dimensions and Functional Gradients

After revealing the relationship between functional gradients and emotional symptoms, we further integrated 17 demographic, lifestyle, and nutritional variables and conducted a partial least squares correlation (PLS-C) analysis. Permutation testing (10,000 permutations) showed that only the first latent variable (LV-1) reached a significant level (cross-block correlation r = 0.25, p = 0.048; Fig. 4A).

**Figure 4.**
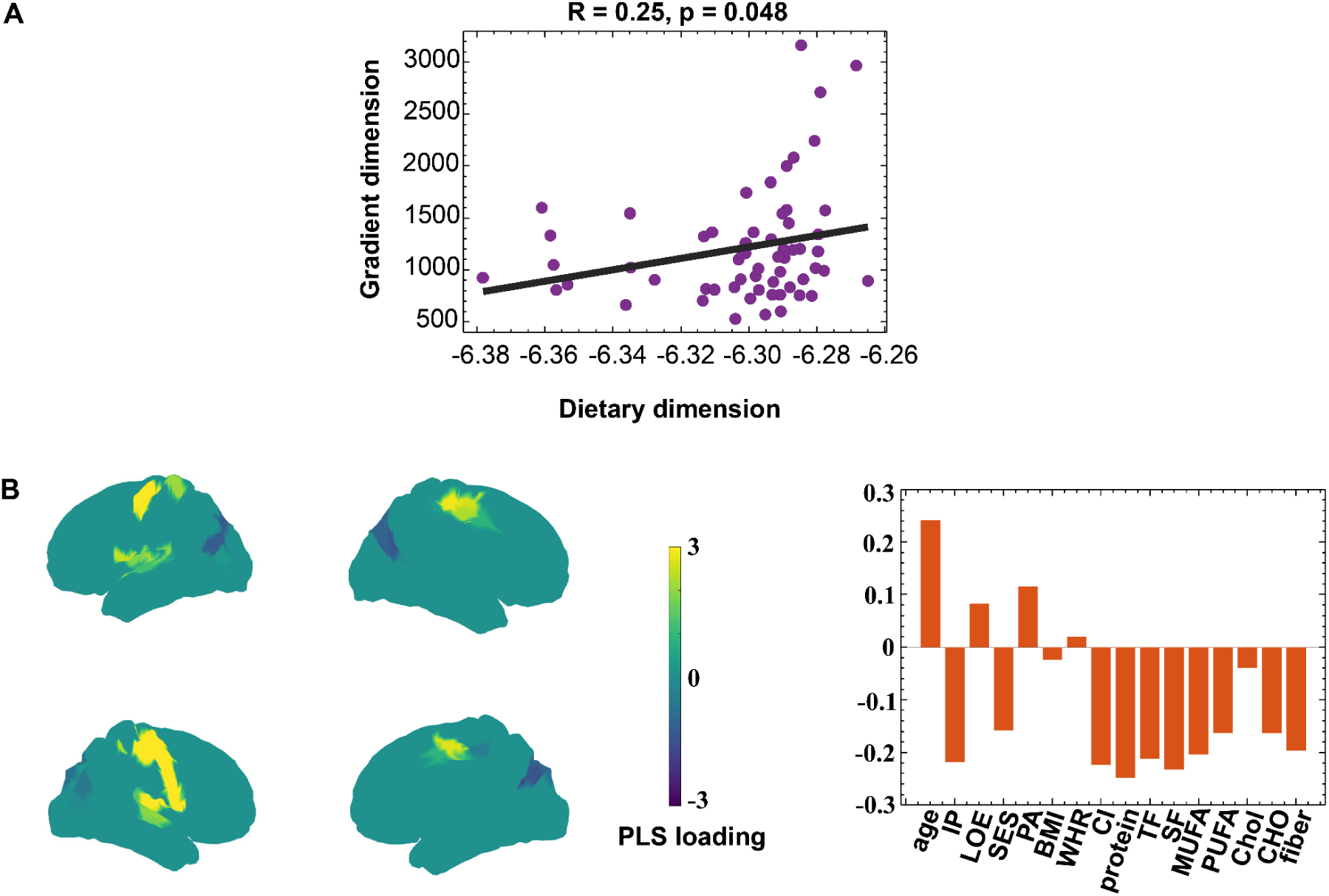
Association between dietary dimension and brain gradient dimension, and PLS loadings. (A) Scatter plot showing the correlation between dietary dimension and brain gradient dimension. The purple dots represent individual data points, and the black line indicates the regression fit. A significant positive correlation was observed (R = 0.25, p = 0.048).(B) Left: Brain regions showing partial least squares (PLS) loadings, with a color bar indicating loading values (ranging from –3 to 3). Right: Loadings of behavioral and dietary variables in the PLS model, including age, intestinal parasite (IP), level of education (LOE), socioeconomic status (SES), physical activity (PA), body mass index (BMI), waist-to-hip ratio (WHR), caloric intake (CI), protein, total fat (TF), monounsaturated fatty acids (MUFA), polyunsaturated fatty acids (PUFA), cholesterol (CHol), carbohydrates (CHO), and dietary fiber.

In terms of brain-side loadings, LV-1 was characterized by increased weights in the central sulcus and dorsal parietal lobe (motor-somatosensory regions) and decreased weights in the visual cortex (Fig. 4B left). On the behavioral side, after 5,000 bootstrap estimations, the main positive loadings of LV-1 included age, educational level, physical activity, and waist-to-hip ratio; negative loadings included intestinal parasite carriage, lower socioeconomic status (SES), lower body mass index (BMI), and intake of all macronutrients (protein, total fat, monounsaturated/polyunsaturated fatty acids, carbohydrates, and dietary fiber; Fig. 4B right).

This result suggests that functional gradient-microbiota coupling is not limited to neural or microbial levels but is embedded in a broader health context. This reveals a quantifiable axis spanning the brain-behavior-microbiota system, which is closely associated with metabolic and lifestyle indicators.

### 2.5 Molecular-Receptor Enrichment and Gene Pathways

Finally, the results of spatial transcriptomic enrichment analysis revealed its molecular basis: significant gene terms were concentrated in Rho-GTPase-mediated cytoskeletal remodeling and GPCR-dependent synaptic signal transduction, accompanied by vesicular transport and integrin adhesion processes (Fig.5A). The receptor profile further showed that transmodal-related brain regions were enriched with GPCR-coupled receptors of multiple neurotransmitter systems (5-hydroxytryptamine [5-HT], dopamine [DA], acetylcholine [ACh], gamma-aminobutyric acid [GABA], glutamate, histamine, cannabinoid, and opioid; Fig.5B). These findings suggest that FR_Rumi_unc may activate the GPCR–Rho/Integrin–vesicle cascade through metabolites such as butyrate, thereby modulating synaptic plasticity and the macroscopic functional hierarchy of the brain.

**Figure 5.**
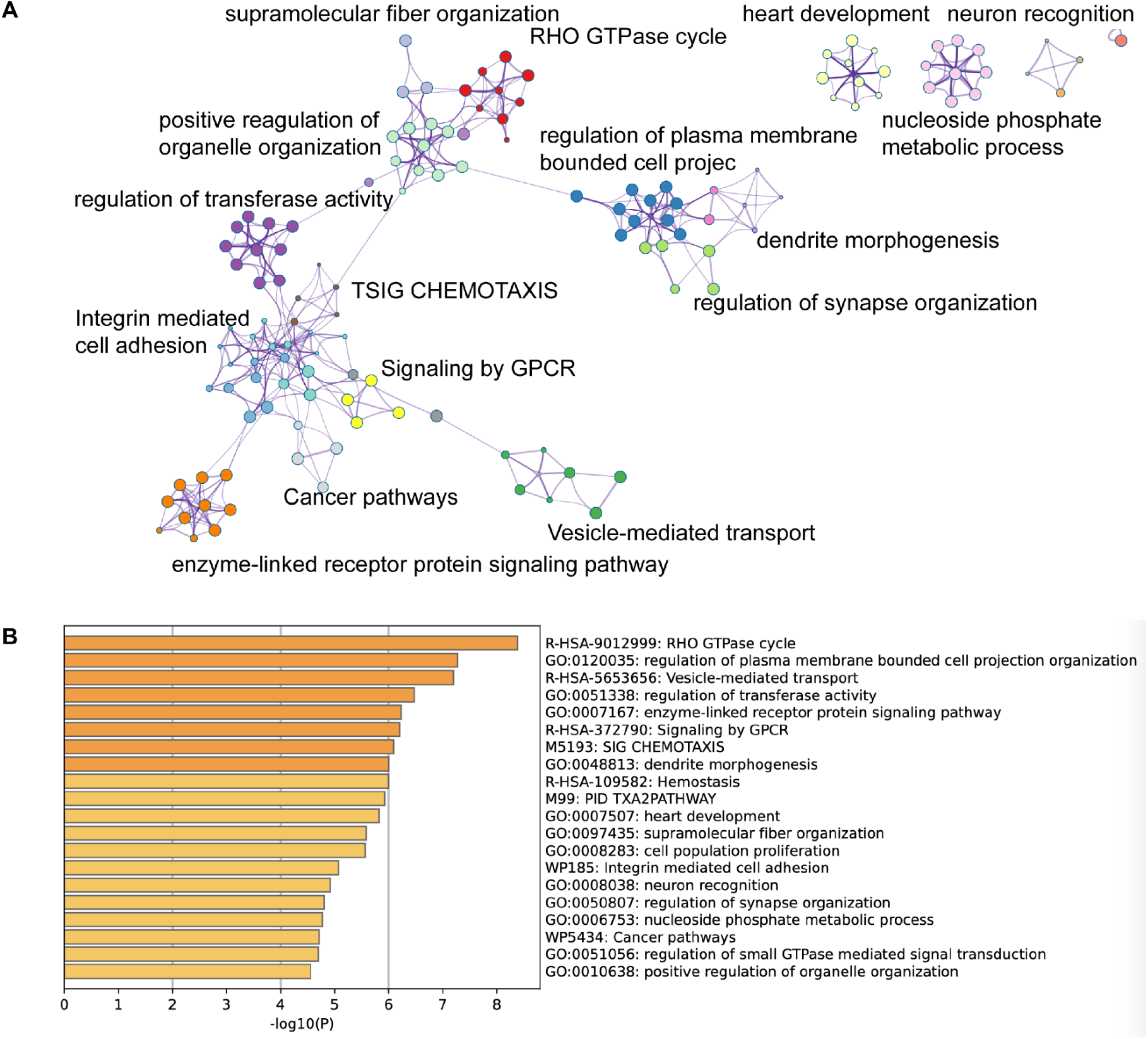
Functional enrichment and pathway analysis of target genes. (A) Network visualization of the significantly enriched biological processes and pathways. Each node represents a functional term, and the edges indicate the genes shared between the terms. Clusters highlight processes including RHO GTPase cycle, integrin-mediated cell adhesion, vesicle-mediated transport, GPCR signaling, chemotaxis, cancer pathways, dendrite morphogenesis, and synapse organization.(B) Bar plot of the top enriched pathways and Gene Ontology (GO) terms ranked by significance (–log10(P)). The list includes pathways such as the RHO GTPase cycle, regulation of plasma membrane-bound cell projection organization, vesicle-mediated transport, transferase activity regulation, signaling by GPCR, chemotaxis, dendrite morphogenesis, heart development, supramolecular fiber organization, cell adhesion, neuron recognition, and cancer pathways.

Overall, these results delineate a multi-level coupling axis of gut microbiota–brain functional gradients–emotional and metabolic health, providing systematic evidence for understanding microbiota-brain-behavior interactions.

## 3. Discussion

Using a cross-modal brain-gut multi-omics dataset, this study systematically delineated the multi-level coupling relationships between the abundance of *Ruminococcaceae_unc* and brain functional gradients, network topology, emotional phenotypes, diet behavior structures, and molecular receptor pathways. Our results demonstrated that the abundance of this specific gut microbiota is not only associated with the hierarchical rearrangement of functional gradients but is also accompanied by network topological drift and is tightly coupled with emotional symptoms and lifestyle factors. At the molecular level, these associations point to the GPCR–Rho/integrin–vesicle cascade pathway. Overall, this multi-scale framework provides new evidence for explaining the microbiota-brain-behavior axis and lays a theoretical foundation for future microbiota-targeted interventions.

First, we found that the abundance of Ruminococcaceae_unc was closely associated with the reweighting of the FG-1. Transmodal hubs, such as the angular gyrus and lateral associative visual cortex, exhibited increased weights, whereas the sensory-motor and gustatory cortices showed decreased weights. This finding extends previous research on the relationship between gut microbiota abundance and functional connectivity, which primarily examined nodes or edges within discrete networks, such as the coupling between autonomic homeostasis and the central executive network. In contrast, our gradient-based approach reveals the localization and directionality of microbiota effects within the continuous hierarchical spectrum of the cerebral cortex, and more intuitively demonstrates a "shift from unimodal to transmodal cortices." This suggests that gut microbiota may influence the global hierarchical organization of the brain by regulating the integrative capacity of hub regions.Further analysis of network small-world properties showed that increased *Ruminococcaceae_unc* abundance was accompanied by a decrease in both clustering coefficient and path length, with the network drifting from a small-world state toward a more randomized topology. Small-worldness is considered the optimal balance of the brain between local specialization and global integration. Existing studies have suggested that conditions such as psychiatric and metabolic disorders are often characterized by reduced small-worldness[12–14].

This study is the first to demonstrate a systematic association between gut microbiota abundance and balance in a healthy population. This finding not only supplements evidence for the topological dimension of the microbiota-brain connection but also suggests that the gut microbiota may alter the outcomes of cognitive and emotional processing by enhancing global efficiency.

At the level of clinical relevance, we found that functional gradient differences were negatively correlated with depression and anxiety scores; specifically, the more severe the emotional symptoms, the more the brain tended toward the unimodal end. This result aligns with previous neuroimaging evidence: patients with depression and anxiety often exhibit reduced functionality in transmodal integration networks, such as the default mode network (DMN) and prefrontal hubs[15–16].Our results further suggest that gut microbiota abundance may serve as a potential biological link between neuroimaging features and emotional vulnerability. In other words, functional gradients are not merely neural-level indicators but may also act as cross-level markers reflecting microbiota-emotion interactions. Partial least squares correlation (PLS-C) analysis revealed that functional gradients were significantly coupled with a latent variable integrating demographic, lifestyle, and nutritional variables. Age, educational level, and physical activity showed positive loadings, whereas nutritional intake and body mass index (BMI), among other factors, showed negative loadings. This result is highly consistent with the "diet-metabolism-microbiota-brain" pathway proposed in previous studies, and our analysis revealed that brain-microbiota coupling does not exist in isolation but is embedded within a broader health context, particularly intertwined with nutritional intake and lifestyle indicators. This provides a foundation for future multimodal interventions, such as the combination of dietary manipulation and behavioral training.

Finally, spatial transcriptomic and receptor enrichment analyses revealed the "GPCR–Rho/Integrin–vesicle" axis as a potential molecular mechanism. This result is consistent with previous transcriptomic-neuroimaging studies that have shown that functional gradients are highly coupled with the spatial distribution of neurotransmitter receptors [17, 18]. We further identified that the relevant brain regions are enriched in multiple classes of GPCR-coupled receptors, including 5-hydroxytryptamine (5-HT), dopamine (DA), acetylcholine (ACh), gamma-aminobutyric acid (GABA), glutamate, and cannabinoids, suggesting that gut-derived metabolites, such as butyrate, may modulate synaptic plasticity and functional hierarchical dynamics through these receptor clusters. This finding provides a molecular explanation for how the gut microbiota influences cognition and emotion and lays a theoretical foundation for the design of drugs or prebiotics targeting specific receptors.

Despite the novel evidence provided by this study regarding the integration of the microbiome, brain functional gradients, and transcriptomic information, several limitations exist. First, the analysis of gene and receptor distribution relied primarily on publicly available population-level transcriptomic atlases rather than on individual-level sequencing data. Consequently, it is unable to reveal fine-grained covarying patterns between gut microbiota and host genes at the individual level.Second, this study adopted a cross-sectional design, lacking longitudinal follow-up and interventional evidence. This makes it difficult to infer the causal relationships and dynamic evolutionary trajectories between the variables of interest.

Third, the functional gradients in this study were derived from resting-state functional magnetic resonance imaging (fMRI) signals. Although resting-state fMRI reflects the macroscopic hierarchical organization of the brain, the lack of task-based paradigms limits its capacity to mechanistically interpret specific cognitive processes such as executive control and emotion regulation. Finally, the limited sample size and restricted sex range (exclusively male participants) may compromise the extrapolatability and generalizability of the results.

## 4. Conclusion

In summary, this study systematically integrated functional gradients, network topology, emotional symptoms, diet-related behavior factors, and molecular mechanisms, revealing the key role of *Ruminococcaceae_unc* in the gut microbiota-brain-behavior axis.

Future studies should combine longitudinal tracking, multimodal task-based paradigms, and individual-level gene transcriptomic sequencing with metabolomics to validate causal relationships, elucidate the roles of specific metabolites, such as butyrate, in the GPCR axis, and explore the regulatory potential of personalized dietary or prebiotic interventions on functional gradient plasticity. These efforts will enable a more comprehensive elucidation of the hierarchical neurobiological mechanisms underlying the microbiota-gut-brain axis and are expected to provide new avenues for developing transmodal intervention strategies for depression, anxiety, and metabolism-related disorders.

## 5. Method

### 5.1 Study Population Selection

This study adopted a cross-sectional design and enrolled 88 healthy Colombian males aged 21–40 years. The exclusion criteria included self-reported medical conditions; current or long-term use of any medications; vegetarian or vegan diets; left-handedness; presence of cardiac devices or other MRI-incompatible implants; and alcohol, tobacco, or substance abuse, as assessed by the Alcohol, Smoking, and Substance Involvement Screening Test (ASSIST) Version 1.1.66. Participants who had recently used antibiotics or probiotics or experienced infectious diseases were required to complete a washout period before enrollment.

All participants underwent a physical examination to obtain anthropometric measurements and blood pressure. Biological sample collection and resting-state functional magnetic resonance imaging (rs-fMRI) were performed within a 4-week period. The study was conducted in accordance with the Declaration of Helsinki (1975) and Resolution No. 8430 of the Colombian Ministry of National Health and was approved by the Research Ethics Committee of the School of Medicine, University of Antioquia (No. 007, May 11, 2017). All participants provided written informed consent.

### 5.2 Imaging Dataset

All participants underwent scanning using an Ingenia 3.0T Philips MRI scanner equipped with a 16-channel phased-array rigid head coil. High-resolution 3D T1-weighted sequence: 175 axial slices, repetition time (TR) = 7807 ms, echo time (TE) = 3593 ms, field of view (FOV) = 256 × 256 mm², matrix size = 256 × 256, voxel size = 1 × 1 × 1 mm³, flip angle (FA) = 8°. Resting-state functional magnetic resonance imaging (rs-fMRI): Total duration of 10 minutes, 300 volumes, 36 slices, TR = 2000 ms, TE = 30 ms, FA = 70°, FOV = 112 × 112 mm², matrix size = 72 × 72, voxel size = 2 × 2 × 3 mm³. The participants were instructed to close their eyes, remain awake, and minimize head movement during the scan.

### 5.3 Data Preprocessing and Functional Connectome

For all datasets, raw DICOM files were converted to the Brain Imaging Data Structure (BIDS) format using HeuDiConv version 0.13.1, and preprocessing of structural and functional data was performed using fMRIPrep version 23.0.2 (implemented based on Nipype version 1.8.6) [19, 20]. This tool automatically generates detailed preprocessing reports, with anatomical preprocessing steps including intensity normalization, brain extraction, tissue segmentation, cortical surface reconstruction, and spatial normalization, while functional preprocessing covers head motion correction, slice-timing correction, and co-registration. Subsequently, the preprocessed functional time series were parcellated according to the Schaefer 400×7 atlas, and confound regression was conducted using Nilearn following the "simple" strategy proposed by Wang et al. [21], which incorporated high-pass filtering, motion regression (with a displacement threshold of 2 mm), tissue signal regression, linear detrending, and z-transformation. Finally, individual-level functional connectivity matrices were calculated using zero-lag Pearson correlation coefficients and standardized using Fisher’s r-to-z transformation.

### 5.4 Functional Organization Gradients

Whole-cortical functional connectome gradients were generated using BrainSpace (version 0.1.10; https://github.com/MICA-MNI/BrainSpace) with the default parameters. After z-transformation of the connectivity data, only the top 10% of weighted connections per brain region were retained to enhance the robustness of the gradient estimation, consistent with previous studies [5]. To quantify the similarity of connectivity patterns across brain regions, an affinity matrix was constructed using the cosine similarity [22]. Subsequently, diffusion map embedding, a nonlinear manifold learning method, was applied to derive a low-dimensional representation of each participant’s high-dimensional functional connectome. Among the resulting gradient components, the top ten were extracted; however, this study focused exclusively on the primary gradient, as it captures the dominant functional axis spanning from the sensory to the association cortices. Finally, individual-level gradient maps were aligned to a template gradient derived from 100 unrelated healthy adults (Human Connectome Project [HCP] data) using Procrustes rotation [23].

In the inferential statistics, we first calculated the FG-1 metric at the parcellation level and conducted correlation modeling at the group level. Specifically, we employed multivariate linear and partial correlation models to examine the association between FG-1 and the relative abundance of target gut microbiota, such as *Ruminococcaceae_unc*, while controlling for covariates, including age, sex, and head motion (mean framewise displacement, mean FD). Multiple comparisons were corrected using the false discovery rate (FDR) method (two-tailed test; q < 0.05). To improve spatial interpretability, the parcellation-level statistical results were projected onto the cortical surface for visualization, with the location and directionality of the significant clusters reported in the figures. For key results, robustness checks were conducted using a more stringent cluster-defining threshold (pFDR < 0.001 at the vertex/parcellation level and cluster-level FDR < 0.05) to ensure that the spatial statistics were not driven by a limited number of units.

### 5.5 Graph Theory Analysis

We extracted two global metrics: (1) the average clustering coefficient, which quantifies the local interconnectivity of brain regions and indicates how tightly neighboring regions form interconnected subgroups, and (2) the average path length, which represents the mean shortest path between any two regions within the network [24, 25]. At the regional level, we adopted a strategy analogous to that used for the global metrics. Specifically, for each cortical and subcortical region in the thresholded structural covariance network, we calculated the normalized clustering coefficient, normalized path length, and their ratios. Based on the distribution of effect sizes, we generated topological feature maps to identify regions that deviated from the small-world organization [26]. For instance, regions exhibiting simultaneous increases in both clustering coefficient and path length were classified as "regularized," while those with decreases in both metrics were deemed "randomized." This approach provides a more intuitive framework for evaluating the structural properties and potential abnormalities of brain networks in multiple dimensions.

### 5.6 Fecal Sample Collection, Bacterial DNA Extraction, and Sequencing

Participants collected fresh fecal samples in sterile plastic containers, which were immediately stored under refrigerated conditions and transported to the laboratory within 2 hours of collection. Upon arrival, total DNA was extracted using the Fecal RNA/DNA Isolation Kit (Norgen Biotec Corp.) according to the manufacturer’s instructions. DNA concentration was measured using a NanoDrop™ 2000 spectrophotometer (Thermo Scientific™), and all DNA samples were uniformly diluted to 10 ng/μL as the starting amount for library construction.Library preparation and sequencing were commissioned to Macrogen Inc. (Seoul, South Korea) using the Illumina MiSeq platform with a 300 bp paired-end sequencing protocol. The V3–V4 hypervariable regions of the 16S rRNA gene (for bacteria and archaea) were amplified using the primers Bakt_341F (5′-CCTACGGGNGGCWGCAG-3′) and Bakt_805R (5′-GACTACHVGGGTATCTAATCC-3′). To be compatible with the sequencing workflow, Illumina adapters and corresponding pad/linker sequences were added to the 5’-ends of both forward and reverse primers.

### 5.7 Taxonomic Bioinformatics Analysis of Gut Microbiota

Sequence processing was performed using Mothur v1.44, following the standard operating procedure (SOP) for MiSeq. The workflow was as follows: First, paired-end reads were assembled into contiguous sequences using the make.contigs command. The resulting sequences were aligned and calibrated against the SILVA 16S rRNA reference database (v138.1). Chimeras were detected and removed using VSEARCH, and non-bacterial sequences were filtered out uniformly. Next, inter-sequence distances were calculated using dist.seqs, and reads were clustered into operational taxonomic units (OTUs) at a 3% dissimilarity threshold (0.03).

At the sample level, normalization was conducted using total-sum scaling (TSS), and rare OTUs with an abundance of <3 reads were removed. Taxonomic annotation was performed using the Ribosomal Database Project (RDP) Classifier with a bootstrap threshold of 80%. The results were exported in BIOM format and imported into the R environment for statistical analysis using the phyloseq and microbiome packages. In R, the median number of sample reads was used as a scaling factor to calculate the normalized counts for each bacterial taxon; based on this, the relative abundance and dominant taxa at the phylum and family levels were obtained. α-diversity indices were estimated using the phyloseq package.

### 5.8 Dietary and Behavioral Assessments

Habitual diet was evaluated using the online version of a locally developed self-reported semi-quantitative food frequency questionnaire (CFIA) [27], which assessed the intake frequency and portion sizes of various foods over the previous year. Internally developed R scripts were used to calculate the daily total energy intake and major macronutrients, including protein, total fat, monounsaturated fatty acids, polyunsaturated fatty acids, carbohydrates, and dietary fiber.

Lifestyle indicators were assessed using the International Physical Activity Questionnaire-Short Form (IPAQ-S). The demographic variables included age, educational level, socioeconomic status (SES), body mass index (BMI), and waist-to-hip ratio (WHR). Emotional status was evaluated using the Zung Self-Rating Depression Scale (with a cutoff value of >50 for depressive symptoms) and the State-Trait Anxiety Inventory (STAI-S >41 for state anxiety; STAI-T >44 for trait anxiety) [28, 29].

### 5.9 Associations of Functional Gradients with Clinical Symptoms, Diet, and Behavior

To examine the clinical relevance of functional gradient alterations, we first extracted the FG-1 weight values of cortical regions that were significant in the correlation analysis with *Ruminococcaceae_unc* abundance. These values were then subjected to Pearson’s correlation analysis with scores from emotional scales (Zung Depression Scale, STAI-S, STAI-T), while controlling for covariates including age, sex, and head motion (mean framewise displacement, mean FD).

Furthermore, to reveal the integrated patterns between functional gradients and dietary/lifestyle variables, partial least squares correlation (PLS-C) analysis was performed. This analysis modeled the association between individual-level FG-1 metrics and 17 variables, including demographics, nutrition, and behavior. Significance was determined via 10,000 permutation tests, and the stability of the loadings was evaluated using 5,000 bootstrap resamplings.

### 5.10 Functional Reorganization and Receptor Mapping

Receptor densities were evaluated using data from positron emission tomography (PET) tracer studies encompassing 18 receptors and transporters across nine neurotransmitter systems. The relevant data were recently shared by Hansen et al. (https://github.com/netneurolab/hansen_receptors)[8]. These neurotransmitter systems include dopamine (D1, D2, DAT), norepinephrine (NET), serotonin (5-HT1A, 5-HT1B, 5-HT2, 5-HT4, 5-HT6, 5-HTT), acetylcholine (α4β2, M1, VAChT), glutamate (mGluR5), γ-aminobutyric acid (GABAa), histamine (H3), cannabinoids (CB1), and opioids (MOR) [30].Voxel-level PET images were aligned to the MNI-ICBM 152 nonlinear 2009 template (version c, asymmetric), and the participant images were averaged across individual studies. Subsequently, the averaged PET images were parcellated according to the Schaefer 200 atlas. For receptors or transporters with multiple averaged images obtained from the same tracer, such as 5-HT1B, D2, and the vesicular acetylcholine transporter (VAChT), a weighted average was calculated for the integration.

In this study, we aimed to explore the topological correlations between functional reorganization patterns and other salient features. To infer these correlations, we implemented a null model that systematically disrupts the relationship between the two topological maps while preserving spatial autocorrelation [31]. First, the receptor maps were randomly shuffled, and their relationship with functional reorganization patterns was tested. Next, the null model was generated by randomly sampling and rotating the spatial coordinates, followed by reassigning the node values based on the nearest result parcellation. This process was repeated 1000 times. Notably, rotation was first applied to one hemisphere and then mirrored onto the other hemisphere. The threshold was determined using the 95th percentile of the shuffling occurrence frequency in the spatial null model.

## DATA AVAILABILITY

The authors have deposited raw gut microbiota sequences into the Sequence Read Archive (SRA) with the BioProject accession number PRJNA1000574. Neuroimages were deposited into the Openneuro repository https://openneuro.org/git/3/ds004648.

## CODE AVAILABILITY

Code will be available on https://github.com/Laoma29/Publication_codes.

## ACKNOWLEDGMENTS

Xiaobo Liu is supported by the China Scholarship Council.

